# The Addiction Risk Factor: A Unitary Genetic Vulnerability Characterizes Substance Use Disorders and Their Associations with Common Correlates

**DOI:** 10.1101/2021.01.26.21250498

**Authors:** Alexander S. Hatoum, Emma C. Johnson, Sarah M.C. Colbert, Renato Polimanti, Hang Zhou, Raymond Walters, Joel Gelernter, Howard J. Edenberg, Ryan Bogdan, Arpana Agrawal

**Affiliations:** Washington University School of Medicine, Department of Psychiatry, Saint Louis, USA; Department of Psychiatry, Division of Human Genetics, Yale School of Medicine, New Haven, CT, USA; Veterans Affairs Connecticut Healthcare System, West Haven, CT, USA; Analytic and Translational Genetics Unit, Department of Medicine, Massachusetts General Hospital and Harvard Medical School, Boston, MA, USA; Stanley Center for Psychiatric Research, Broad Institute of MIT and Harvard, Cambridge, MA, USA; Department of Genetics, Yale School of Medicine, New Haven, CT, USA; Department of Neuroscience, Yale School of Medicine, New Haven, CT, USA; Department of Medical and Molecular Genetics, Indiana University School of Medicine, Indianapolis, IN, USA; Department of Biochemistry and Molecular Biology, Indiana University School of Medicine, Indianapolis, IN, USA; Department of Psychological & Brain Sciences, Washington University in St. Louis

## Abstract

Substance use disorders commonly co-occur with one another and with other psychiatric disorders. They share common features including high impulsivity, negative affect, and lower executive function. We tested whether a common genetic factor undergirds liability to problematic alcohol use (PAU), problematic tobacco use (PTU), cannabis use disorder (CUD), and opioid use disorder (OUD) by applying genomic structural equation modelling to genome-wide association study summary statistics for individuals of European ancestry (Total N = **1**,**019**,**521**; substance specific Ns range: **82**,**707-435**,**563**), while adjusting for the genetics of substance use (Ns = **184**,**765-632**,**802**). We also tested whether shared liability across SUDs is associated with behavioral constructs (risk taking, executive function, neuroticism; Ns = **328**,**339-427**,**037**) and non-substance use psychopathology (psychotic, compulsive, and early neurodevelopmental disorders). Shared genetic liability to PAU, PTU, CUD, and OUD was characterized by a unidimensional addiction risk factor (termed *The Addiction-Risk-Factor*, independent of substance use. OUD and CUD demonstrated the largest loadings, while problematic tobacco use showed the lowest loading. *The Addiction-Risk-Factor* was associated with risk taking, neuroticism, executive function, and non-substance psychopathology, but retained specific variance before and after accounting for genetics of substance use. Thus, a common genetic factor partly explains susceptibility for alcohol, tobacco, cannabis, and opioid use disorder. *The Addiction-Risk-Factor* has a unique genetic architecture that is not shared with normative substance use or non-substance psychopathology, suggesting that addiction is not the linear combination of substance use and psychopathology.

## INTRODUCTION

Substance use and use disorders (SUDs) represent large and growing public health problems that account for nearly 6% of global disease burden^1^. SUDs, both licit and illicit, commonly co-occur with each other and also with non-substance psychopathology; comorbidity is associated with increased symptom severity^2^ and worse outcomes (e.g., less responsivity to treatment, greater socioeconomic costs^3^). However, the etiology underlying shared risk across these disorders is poorly understood.

### Shared Genetic Liability

According to twin studies, the moderate-large heritability (50-60%) of distinct SUDs (i.e., alcohol, nicotine, cannabis, and other illicit drugs) is partly attributable to a shared genetic vulnerability.^4^ Similarly, genetic correlations estimated from genome-wide association study (GWAS) data support a shared genetic vulnerability between SUDs (e.g., SNP-rG = .73 between alcohol use disorder and opioid use disorder),^5^ between SUDs and substance use (e.g., SNP-rG = .78 between problematic alcohol use and drinks per week),^6^ and between SUDs and psychopathology (e.g., SNP-rG = .33 between cannabis use disorder and major depressive disorder^7^). What remains unclear is the extent to which genetic liability across substance use *disorders* is *shared with* and *distinct* from that of substance *use* (but not dependence) and non-substance psychopathology, and what putative intermediate phenotypes may link shared genetic liability between SUDs and non-substance psychopathology.

#### Substance Use and Use Disorder

Substance use and SUDs have substantial genetic overlap; however, genetic mechanisms that relate to SUD liability beyond normative or frequently occurring substance use remain. For opioids^8^, alcohol^9,10,11^, and cannabis^7^, the use and use disorder dimensions show differing associations with psychopathology (e.g. schizophrenia) and life outcomes (e.g. educational attainment)^7,8,12,13^.

#### Substance use and Psychopathology

Recently, Lee and colleagues^14^ identified 3 broad clusters (psychotic, compulsive, and early neurodevelopmental) representing shared and distinct genetic liability to 8 non-substance psychiatric disorders. While there has been limited integration of substance use phenotypes into psychopathology models, polygenic liability to cross-diagnostic vulnerability is associated with general substance use and SUDs^15^. Further, emerging evidence suggests partial overlap: tobacco *use* shares variance with ADHD^16^, alcohol and cannabis dependence load with antisocial behavior^17^, and alcohol use and use disorder load together onto an Externalizing factor^18^. Collectively, these data suggest that substance use and use disorders share some common genetic liability with psychopathology.

#### Stage-Based Addiction Individual Differences and Substance Use Disorders

SUD vulnerability has been conceptualized within a 3-stage neurobiological model consisting of binge/intoxication, preoccupation/anticipation, and withdrawal/negative affect^19^. In this model, initial positive reinforcement is derived from stimulation of neural reward circuitry that drives impulsive behaviors in the context of under-developed tolerance. With continued use and progression towards SUD, the reinforcing properties of substances shift from positive to negative reinforcement; as use becomes compulsive, it functions to return the body to drug-present homeostasis and alleviate low mood, a predisposition to which is broadly indexed by neuroticism^20^. Following repeated drug-reward and drug-homeostasis pairings, cognitive preoccupation with the drug in expectation of reward/relief emerges in the context of impaired executive functioning^21^. While GWASs support genetic correlations between SUDs and risk-taking^5,22^, Executive Functioning^23^, and negative affect^5,22^, the extent to which common genetic liability across SUDs relates to these constructs has yet to be examined.

### The Current Study

Given evidence of shared liability to SUDs and other forms of psychopathology, understanding the shared and unique genetic contributions to SUDs and how these relate to heritable proxies for stage-based addiction constructs, non-substance psychopathology, and substance use may generate etiologic insights that improve psychiatric nosology, prevention, and treatment. To this end, we *first* estimate the shared genetic structure across SUDs by applying genomic structural equation modelling (gSEM)^24^ to summary statistics generated by the largest GWAS of problematic alcohol use (PAU)^22^, problematic tobacco use (PTU)^25,26^, cannabis use disorder (CUD)^7^, and opioid use disorder (OUD)^5^. We name the shared variance across SUDs the *Addiction-Risk-Factor. Second*, we relate the *Addiction-Risk-Factor* to genetics of behavioral constructs representing proxies of the stage-based model of SUDs. We estimate the extent to which genetic liability to risk-taking, executive function, and neuroticism are related to *The Addiction-Risk-Factor. Third*, we examine whether *The Addiction-Risk-Factor* is associated with the 3 factors representing genetic liability to non-substance psychopathology^14^ (i.e., psychotic, compulsive, and neurodevelopment) and whether stage-based addiction constructs (i.e., risk-taking, executive function, neuroticism) indirectly link *The Addiction-Risk-Factor* to psychopathology. *Finally*, given that genetic liability to substance use (e.g., ever using, quantity-frequency) and later stages of SUDs are partially distinct^7,12,13^, we repeat all analyses while incorporating genetic liability to substance use (i.e., alcohol drinks/week^26^; tobacco ever regularly use^26^, cannabis ever use^27^) as covariates.

We hypothesized that SUDs and problem substance use would be largely characterized by a common genetic vulnerability (i.e., *The Addiction-Risk-Factor*) with evidence of potentially important substance-specific liability (e.g., metabolic and signaling pathways for a specific drug such as *ADH1B* variants with alcohol^28^). We hypothesized that (i) *The Addiction-Risk-Factor* would be associated with all 3 non-substance psychiatric clusters while retaining variance unique to itself, (ii) genetic liability to behavioral phenotypes representing vulnerability to stage-based addiction constructs (i.e., risk-taking, executive function, and neuroticism) would be associated with *The Addiction-Risk-Factor* and account for a proportion of the association between *The Addiction-Risk-Factor* and psychopathology factors, and (iii) after accounting for genetics of substance use, *The Addiction-Risk-Factor* would retain unique variance (i.e., we expect significant residual genetic correlations among SUDs) and maintain similar patterns with non-substance psychopathology and stage-based constructs.

## METHODS

### Samples

Summary statistics from the largest available discovery GWASs were used to represent genetic risk for each construct (more details are in **Supplemental Table 1**). These include: **i) 4 *SUDs*** (problematic alcohol use^22^, problematic tobacco use^25^, cannabis use disorder^7^, opioid use disorder^5^); **ii)** 3 ***substance use phenotypes*** (alcohol drinks/week^26^, lifetime ever smoking^26^, lifetime cannabis use^27^); **iii)** 3 ***traits proxying the stage-based model of SUDs*** (risk-taking, executive function, neuroticism); and **iv) 9 *non-substance psychiatric disorders***. Analyses were restricted to data from individuals of European ancestry because GWAS on these constructs in other ancestral origins are not available or are underpowered, and cross-ancestry analysis can confound genetic correlation estimates^29^. All GWAS summary statistics were filtered to retain variants with minor allele frequencies > 0.01 and INFO score > 0.90 for GSCAN and PGC^7,26^ and INFO score > 0.70 for the MVP^5,30^.

**Table 1.** Behavioral Liabilities Mediate the Association Between *The Addiction-Risk-Factor* and Psychiatric Factors. Indirect associations from a mediation model (see Figure 4) where stage-based constructs link non-substance psychopathology (3 factors from Lee et al.,) and *The Addiction-Risk-Factor*. F1 = compulsive disorders, F2 = Psychotic disorders, F3 = Neurodevelopmental disorders. The proportion accounted for by the indirect association (%) and the significance of the indirect association are shown. * P < .05 for the indirect association pathway.

|  | **F1 Indirect %** | **F1 Indirect P** | **F2 Indirect %** | **F2 Indirect P** | **F3 Indirect %** | **F3 Indirect P** |
| --- | --- | --- | --- | --- | --- | --- |
| **No covariates** |  |  |  |  |  |  |
| executive function | 0.007 | 0.463 | **0.051\*** | **0.001\*** | 0.003 | 0.486 |
| Neuroticism | 0.005 | 0.842 | 0.003 | 0.836 | 0.013 | 0.777 |
| Risk-taking | **-0.090\*** | **0.001\*** | **0.152\*** | **1.47E-08\*** | **0.043\*** | **0.013\*** |
| **Controlling for Genetics of Substance Use** |  |  |  |  |  |  |
| executive function | 0.013 | 0.233 | **0.048\*** | **0.047\*** | -0.007 | 0.491 |
| Neuroticism | 0.011 | 0.687 | -0.022 | 0.693 | -0.040 | 0.689 |
| Risk-taking | 0.001 | 0.509 | 0.019 | 0.446 | -0.002 | 0.741 |

### Problematic Substance Use/Substance Use Disorder Summary Statistics

#### Problematic Alcohol Use

Summary statistics for problematic alcohol use (PAU) were derived from a meta-analysis of GWASs of DSM-IV alcohol dependence from the Psychiatric Genomics Consortium^11^ (PGC-AD; n = 11,569 case, 34,999 controls), ICD-9/10 based diagnoses of alcohol use disorders from the Million Veteran Program phase 1 and 2 data (MVP; n = 45,995 cases; 221,396 controls)^9^ and the Problem subscale score from the Alcohol Use Disorders Identification Test (AUDIT-P)^13^ from the UK Biobank (n = 121,604)^22^. The final GWAS summary statistics included data on 435,563 participants^22^. We also report on model fit with PGC-AD (instead of PAU) in the supplement (**Alternative Models, M1)**.

#### Problematic Tobacco Use (PTU)

We used summary statistics from the GWAS of the Fagerstr*ö*m Test for Nicotine Dependence^25^ (FTND). As cigarettes per day is an item within the FTND and the genetic correlation between FTND and cigarettes per day is high (calculated rG = 0.97 CI = .12)^12^, we combined CPD And FTND into a single indicator. We applied Multi-Trait Analysis of Genome-wide association study summary statistics (MTAG^31^) to summary statistics generated from the GWAS and Sequencing Consortium of Alcohol and Nicotine Use (GSCAN) GWAS of cigarettes per day to create the combined problematic tobacco use (PTU) phenotype^26^. The final GWAS summary statistics had an effective sample size of n = 270,120 individuals. We also report on model fit with just FTND as an indicator in the supplement (**Alternative Models M2)**.

#### Cannabis Use Disorder (CUD)

Summary statistics were derived from a GWAS meta-analysis^7^ of DSM-IV and DSM-III-R cannabis abuse and dependence from the Psychiatric Genomics Consortium (n = 5,289 cases; n = 10,004 controls), ICD-10 cannabis use disorder from the Lundbeck Foundation Initiative for Integrative Psychiatric Research (iPSYCH) (n = 2,758 cases; n = 53,326 controls), and hospital-based diagnoses from deCODE (n = 6,033 cases; n = 280,396 controls). The final European-ancestry sample included 14,080 cases with CUD and 343,726 controls.

#### Opioid Use Disorder

Opioid use disorder (OUD): Summary statistics were derived from a meta-analysis^5^ of GWASs of DSM-IV opioid abuse or dependence from Yale-Penn, and the Study of Addiction: Genetics and Environment, and ICD-9/10 codes for opioid use disorder from the Million Veteran Program (n = 10,544 cases; n = 72,163 opioid-exposed controls).

### Substance Use Summary Statistics

#### Alcohol use

Alcohol use summary statistics were derived from the GSCAN GWAS^32^ for current (this past week or average in the past year) reported drinks/week (n = 537,349). There was a strong correlation with lifetime PAU (SNP-rG between drinks/week and PAU = 0.77±0.02)^22^.

#### Lifetime Tobacco Use

Summary statistics came from the GSCAN GWAS of reported ever/never regular cigarette smoking (ever n = 301,524, never n = 331,278). There was a moderate correlation with PTU (SNP-rG = 0.28±0.03).

#### Lifetime Cannabis Use

We used summary statistics from a meta-analysis of lifetime cannabis ever-use from the International Cannabis Consortium and UK Biobank (ever n = 43,380; never n = 118,702)^27^. There was a moderate correlation with CUD^7^ (SNP-rG = 0.47±0.05).

### Stage-based Behavioral Constructs

The three-stage behavioral model of addiction focuses on “state” changes in substance use behaviors. Because GWASs measure individual differences in traits, we selected behaviors that (1) are known to convey vulnerability to each stage as proxies, and (2) are heritable.

#### Risk-taking and sensitivity to reward

A GWAS of risk-taking derived from a single item in the UK Biobank (“Would you describe yourself as someone who takes risks?”; data field #2040; risk taker n = 83,677; non-risk taker n = 244,662)^33^.

#### Executive Function

The **“**preoccupation/anticipation” stage is characterized by maladaptive reward valuation and future planning. Recent work argues that this vulnerability is captured by executive functioning^34^. Summary statistics from a GWAS of a latent factor representing common executive functioning were used (N = 427,037)^23^.

#### Negative Emotionality and Sensitivity to Stress

The stage of withdrawal/negative affect represents substance use functioning to mitigate aversive withdrawal symptoms, such as negative affect. Neuroticism has been found to modify stress sensitivity and neural reward processing^35^. Neuroticism was chosen as a trait-based measure representing liability to negative affect as opposed to depression because depression was included in the non-substance psychiatric disorder factor generation and because neuroticism includes trans-diagnostic constructs such as negative urgency (i.e., impulsive attempts to cope with negative affect) that may place individuals at risk for the negative reinforcing aspects of SUDs. We selected the largest GWAS of neuroticism as a heritable proxy (N = 390,278)^20^.

### Non-Substance Summary Statistics

Summary statistics from the PGC Cross-disorder GWAS on the 8 disorders that were previously shown to fit a 3 factor confirmatory model were used^14^. These disorders included Schizophrenia^36^, Bipolar disorder^37^, Major depressive disorder^38^, Attention Deficit Hyperactivity disorder^39^, Obsessive Compulsive Disorder^40^, Anorexia Nervosa^41^, Tourette Syndrome^42^, and Autism Spectrum Disorder^43^ (See **Supplemental Table 1 for details**).

### Statistical analysis

*First*, we estimated the pairwise genetic correlations between PAU, PTU, CUD and OUD using Linkage Disequilibrium Score Regression (LDSR)^29^. After confirming that the four SUDs were significantly genetically correlated (see **Results**), we applied confirmatory factor analysis to the covariance matrix generated by LDSR using gSEM^44^ with weighted least squares estimation; PAU, PTU, OUD, and CUD indicators were allowed to load freely on a single latent factor (i.e., *The Addiction-Risk-Factor*). Variance of this common latent factor was scaled to 1.0. A residual correlation between PAU and OUD was also estimated to account for measurement overlap, because the Million Veterans Project sample was contained in both PAU and OUD GWAS (but see model fit without this residual correlation in the supplement – **Alternative Models M1**). In supplemental analyses, we also examined alternative two factor models (**Alternative Models M3**).

*Second*, we used a series of structural regression models to estimate the extent to which genetic liability to stage-based constructs of addiction (i.e., risk-taking, executive function and neuroticism) are related to *The Addiction-Risk-Factor*. Here, the *Addiction-Risk-Factor* variance was freed, and the OUD loading was set to 1.0 to scale the model. Intercorrelations were estimated between risk-taking, executive function and neuroticism.

*Third*, we recreated the three factors from Lee et al^14^. (i.e., psychotic disorders, compulsive disorders, and early neuro-developmental disorders) and estimated their relationship with *The Addiction-Risk-Factor* while allowing for inter-factor correlations (the association between *The Addiction-Risk-Factor* and an alternative cross-disorder genetic model from a preprint^45^ was also estimated; this alternative model is shown in **Supplemental Figure 5**). This allowed us to estimate the unique association between each of the 3 psychopathology factors and *The Addiction-Risk-Factor* and to estimate variance that was residual to *The Addiction-Risk-Factor*. We then examined whether proxies for stage-based addiction constructs (i.e., risk-taking, executive function and neuroticism) indirectly linked *The Addiction-Risk-Factor* to the 3 non-substance psychopathology factors using a multiple mediator model. We also conducted supplemental modified Q-Trait analyses^45^ to examine the extent of the mediation (**Q-Trait Analysis**). To estimate residual associations (i.e., direct paths) between the stage-based constructs and *The Addiction-Risk-Factor*, we re-structured the mediation model to one in which the 3 non-SUD psychopathology factors served as “mediators” of the relationship between risk-taking, executive functioning, neuroticism, and *The Addiction-Risk-Factor*.

To separate the genetics of SUD from the genetics of substance use, we estimated models where substance *use* GWAS summary statistics were endogenous predictors of all measured variables in the model. For example, in the model estimating the association between *The Addiction-Risk-Factor* and psychiatric factors, the 8 psychiatric disorders and the 4 SUD disorder variables were regressed on the 3 substance use variables. In this way, covariate effects were estimated simultaneously to our associations of interest.

## RESULTS

### The Addiction Risk Factor

Genetic correlations between problematic alcohol use (PAU^22^), problematic tobacco use (PTU^25,26^), cannabis use disorder (CUD^7^), and opioid use disorder (OUD^5^) ranged from 0.19 (S.E. = .04) to 0.78 (.09) (**Supplemental Figure 1** and **Supplemental Figure 2**). PTU showed the lowest SNP-rG with other SUD phenotypes [i.e., PAU = 0.19 (.04), CUD = 0.31 (.05), OUD = 0.26 (.08)] while OUD showed the highest [PAU = 0.69 (.07), CUD = 0.78 (.09)]. A confirmatory factor model specifying a unidimensional *Addiction-Risk-Factor* underlying the genetic covariance among PAU, PTU, CUD and OUD fit the data well [*X*^2^(1) = .017, p = .895, CFI = 1, SRMR = .002; residual r = .51, p = 0.016; **Figure 1A**]. Loadings were uniformly high except for PTU. Neither PAU nor PTU were impacted by the inclusion of non-diagnostic indices of addiction risk (**Supplemental Results Alternative Models M1-M2**). Alternative 2-factor models did not fit the data well (**Alternative Models M3**).

**Figure 1.**
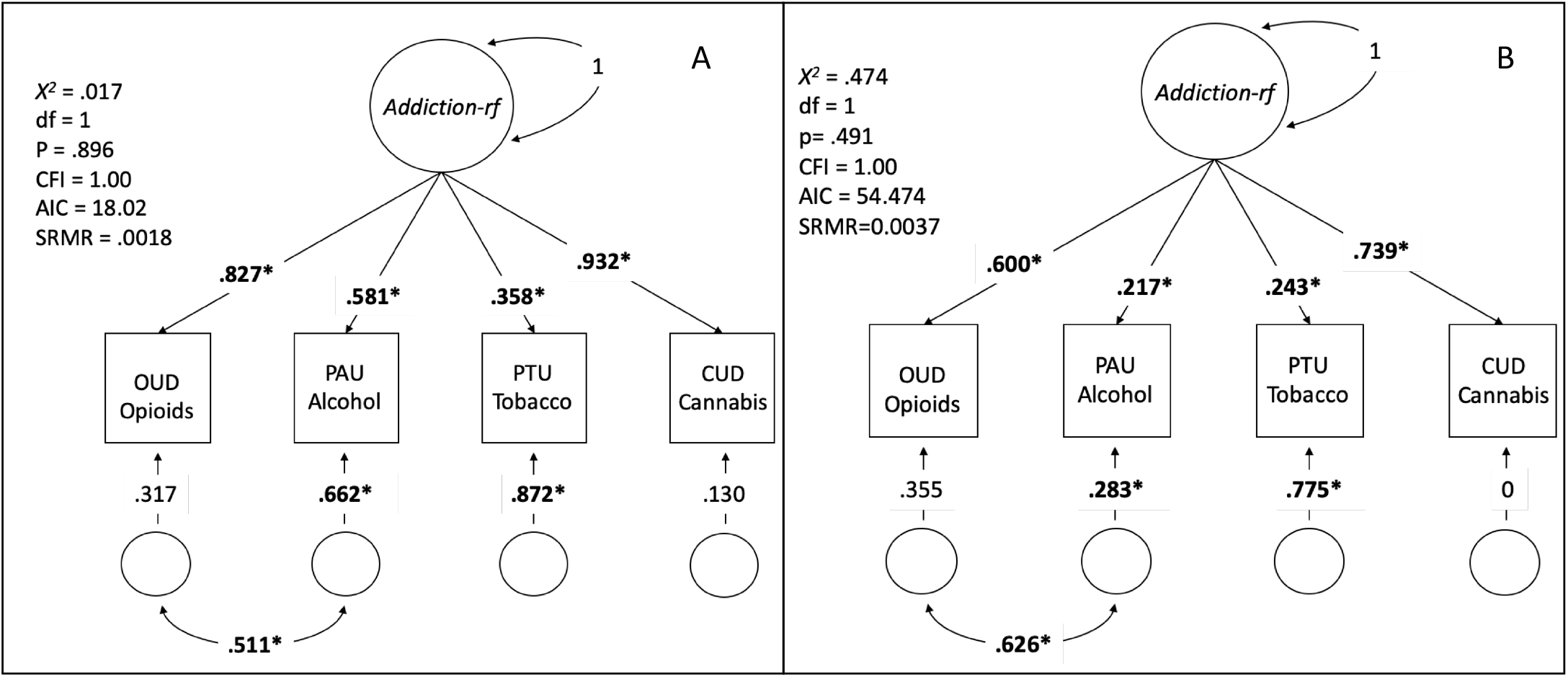
Factor Structure of 4 SUD GWAS. Panel **A**: the model, loadings, and fit for a model that allowed all 4 SUD categories to load on a latent factor. A residual correlation was added between OUD and PAU to account for large sample overlap (both used the Million Veterans Project data; models without residual correlations also fit well: **Supplemental Figure 3**). *Addiction-rf=The Addiction Risk-Factor*. **Panel B**: the same model, but accounting for common substance use (ever smoke, ever use marijuana, and drinks per week) as covariates at the indicator level, i.e. the three substance use measures are exogenous to all indicators in this model and the model represents the residual associations after accounting for substance use. Both models provided excellent fit to the data. **Bold\*** represents significance at p < .05. Note that in panel B, the residual of CUD is zero; this model constraint was necessary, as the model produced a negative residual without the constraint. Note: If you want to recreate the correlation matrix from both panels, the model with residual correlations cannot recover the implied correlation between PAU and OUD without taking the square root of the residual variance, rather than the value of the residual variance itself.

**Figure 2.**
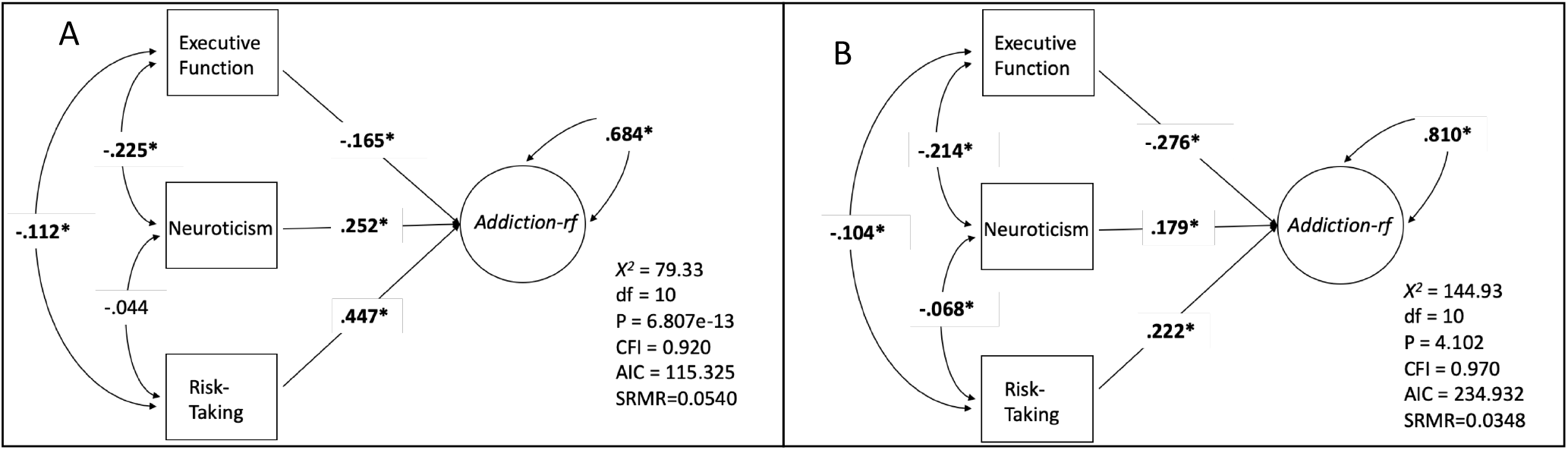
Genetic associations between *The Addiction-Risk-Factor* and behavioral traits: Executive Function, Neuroticism and Risk-taking. Panel A the model, fit, and regression pathways without accounting for common substance use. Panel B is the same model, but accounting for common substance use (ever smoke, ever use marijuana, and drinks per week) as covariates at the indicator level (regressed on all measured variables/GWAS). **Bold\*** represents significance at p < .05.

The inclusion of genetic liability to typical substance use did not modify the single factor structure of *The Addiction-Risk-Factor* (**Figure 1B**); all SUDs continued to load significantly on the factor. However, factor loadings were lower for all substances, especially for PAU, which may be attributable to the high genetic correlation between drinks/week and PAU. Alternative parameterization of substance use as covariates did not improve model fit (**Alternative Models M4**).

### Shared Liability to Stage-based Behavioral Phenotypes

Genetic liability to stage-based addiction constructs was shared with *The Addiction-Risk-Factor* (**Figures 2, Q-Trait Analysis in Supplemental Methods**). As expected, *The Addiction-Risk-Factor* was positively associated with genetic liability to risk-taking (*β* = 0.45) and neuroticism (*β* = 0.25), and negatively associated with executive function (*β* = -0.17; **Figure 2A**). Despite significant genetic overlap between *The Addiction-Risk-Factor* and stage-based behavioral phenotypes, *The Addiction-Risk-Factor* retained unique variance (*Addiction-Risk-Factor* residual = 0.68). When conditioning for genetic liability for substance use, *The Addiction-Risk-Factor* remained significantly associated with increased genetic liability to risk-taking (*β* = 0.22) and neuroticism (*β* = 0.18) and decreased genetic liability to executive function (*β* = -0.28; **Figure 2B**). Accounting for genetic liability for substance use substantially reduced the association between *The Addiction-Risk-Factor* and risk-taking from 0.45 to 0.22 (p_df = 1_ = 4e-09) and accentuated the negative association with executive function from *β* = -0.17 to -0.28 (p_(df = 1)_ = 0.013); there was a smaller effect on the association with neuroticism (from *β* = 0.25 to 0.18, p_(df = 1)_ = 0.012).

### Shared Liability to Non-substance Psychopathology

Genetic liability to non-substance psychopathology (i.e., compulsive disorders, psychotic disorders, and neurodevelopmental disorders) was shared with *The Addiction-Risk-Factor* (**Figure 3**, full models in **Supplemental Figure 4; Supplemental Figure 5** shows results with an alternative cross-disorder model from a recent preprint^45^). Psychotic disorders (*β* = 0.45) and neurodevelopmental disorders (*β* = 0.74) were positively associated with *The Addiction-Risk-Factor* while compulsive disorders showed a negative association (*β* = -0.32; **Figure 3A**). Due to the strong correlation between *The Addiction-Risk-Factor* and early-onset neurodevelopmental disorders (which includes ADHD) we allowed ADHD to load on *The Addiction-Risk-Factor* to control for ADHD; here, an association between *The Addiction-Risk-Factor* and early-onset neurodevelopmental disorders remained, but was significantly attenuated (from *β* = 0.74 to 0.43, p_(df = 1)_ = 5e-5). When conditioning *The Addiction-Risk-Factor* for substance *use*, the psychotic and early neurodevelopmental disorder factors remained significantly associated with *The Addiction-Risk-Factor* (**Figure 3B**). Despite the significant genetic overlap with other psychiatric disorder domains, *The Addiction-Risk-Factor* retained unique variance representing genetic liability specific to SUDs (*The Addiction-Risk-Factor* residual = 0.30, p = 4.54e-3). This unique variance remained significant when accounting for genetic liability to substance use (*The Addiction-Risk-Factor* residual = 0.58, p = 0.015).

**Figure 3.**
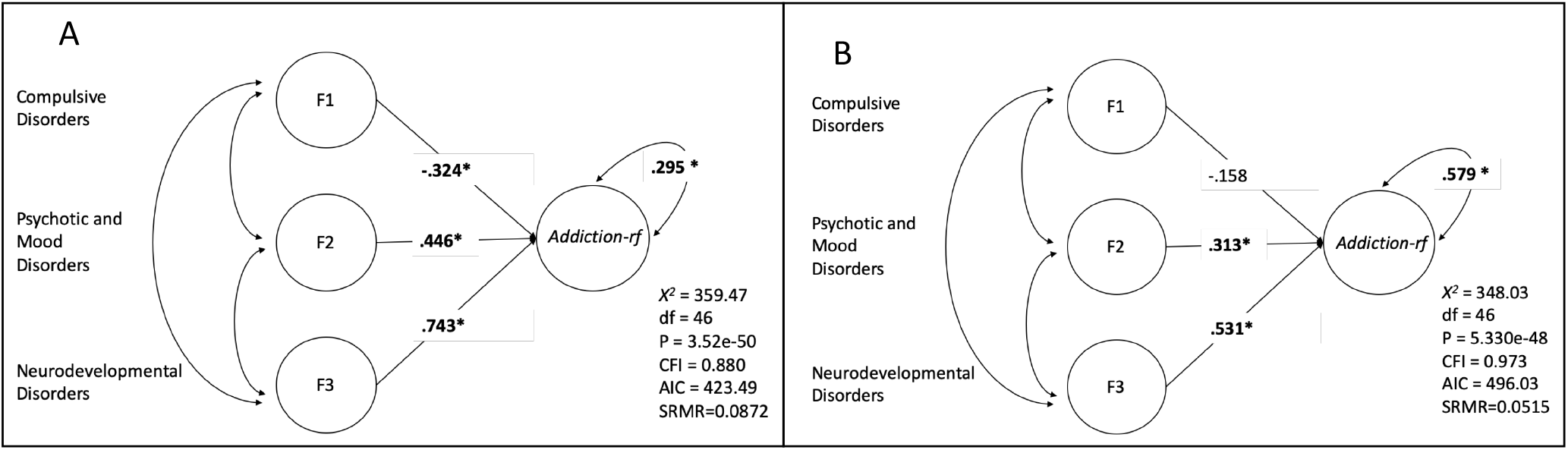
Genetic associations between *The Addiction-Risk-Factor* and Latent Psychopathology Factors: Compulsive disorders (F1; Tourette’s syndrome, Obsessive compulsive disorder, and Eating Disorders), Psychotic Disorders (F2; Major Depressive Disorder, Schizophrenia, and Bipolar Disorder) and neurodevelopmental dysfunction (F3; ADHD, Autism, and Major Depressive Disorder). Panel A the model, fit, and regression pathways without accounting for common substance use (model was scaled by setting the Opioid Use Disorder loading to 1). Panel B is the same model, but accounting for common substance use (ever smoke, ever use marijuana, and drinks per week) as covariates at the indicator level (regressed on all measured variables/GWAS), i.e. the three substance use measures are exogenous to all indicators in this model and the model is the residual associations after accounting for substance use. **Bold\*** represents significance at p < .05. *Addiction-rf = The Addiction-Risk-Factor*.

The specifications for the mediation models are shown **in Supplemental Figure 6**. Genetic liability to risk taking accounted for a proportion of the associations between all non-substance psychopathology domains and *The Addiction-Risk-Factor* (**Table 1**). Executive function uniquely indexed an indirect effect between psychotic disorders and *The Addiction-Risk-Factor* (**Table 1**). When conditioning *The Addiction-Risk-Factor* for genetic liability to substance use, risk-taking no longer accounted for a portion of the association between any non-substance psychopathology domain and *The Addiction-Risk-Factor*, but executive function continued to account for a proportion of the overlap (indirect effect of 0.048) between psychotic disorders and *The Addiction-Risk-Factor* (**Table 1**). *Post-hoc* analyses revealed that executive function retained a unique association with *The Addiction-Risk-Factor* after accounting for genetic liability to both substance use and non-substance psychopathology (**Supplementary Table 2**).

## DISCUSSION

We applied genomic structural equation modeling (gSEM)^24^ to GWAS summary statistics to characterize the genetic influences shared across SUDs and estimate how common genetic liability is related to trait conceptualizations of a theoretical stage-based SUD model as well as to non-substance psychopathology. Three primary findings emerged. *First*, genetic risk for specific SUD phenotypes (i.e., PAU^22^, PTU^25,26^, CUD^7^, and OUD^5^) was largely attributable to a single Addiction risk factor, *The Addiction-Risk-Factor* (**Figure 1**). *Second, The Addiction-Risk-Factor* was associated with genetic liability to trait representations of stage-based facets of addiction (risk taking [binge/intoxication], executive function [preoccupation/anticipation], neuroticism [negative affect]^19^; **Figure 2**). It was also associated with non-substance psychopathology factors (compulsive disorders, psychotic disorders, neurodevelopmental disorders; **Figure 3**). Trait representations of stage-based facets of addiction partially accounted for the shared genetic liability between non-substance psychopathology and *The Addiction-Risk-Factor. Third*, associations between *The Addiction-Risk-Factor* and stage-based constructs and non-substance psychopathology were largely independent of genetic liability to substance *use* phenotypes (i.e., tobacco use, cannabis use, alcoholic drinks/week). However, consistent with the stage-based model of addiction, accounting for substance use attenuated associations between risk taking and *The Addiction-Risk-Factor* while potentiating associations with executive functioning. Collectively, our findings suggest that SUDs are characterized by a common genetic factor, *Addiction-Risk-Factor*.

### *The Addiction-Risk-Factor* retains variance that is not shared with other psychopathology

After accounting for genetic liability to substance use as well as the commonality between *The Addiction-Risk-Factor* and non-substance psychopathology, *The Addiction-Risk-Factor* retained significant variance. These data suggest that *The Addiction-Risk-Factor* may be characterized by unique pathways not shared with substance use or non-substance psychopathology, i.e., addiction is not the linear combination of substance use and psychopathology.

A single latent factor, fit these data well, but specific SUDs showed varying degrees of association. The illicit SUDs (CUD and OUD; **Figure 1**) were almost entirely captured by the common latent factor. Notably, the loading for PTU on *The Addiction-Risk-Factor* was the smallest. One potential contributor to the residual variance of PTU may be the use of FTND and cigarettes/day as indices of PTU. Unlike the Diagnostic and Statistical Manual (DSM) criteria which index psychological and physiological aspects of tobacco use disorder, the FTND is an index of biochemical dependence and although used widely by investigators to define addiction, phenotypically shows only moderate agreement with DSM-defined nicotine dependence (r = 0.50; kappa = 0.3)^46^.

### Proxies of Stage-based Behavioral Constructs and *The Addiction-Risk-Factor*

Behavioral stage-based models of SUD posit a cyclical relationship between positive reinforcement, negative reinforcement, and incentive salience^19^ that we found can be (partially) captured by genetic liability to risk-taking, executive functioning, and negative emotionality (neuroticism). The strongest association with *The Addiction-Risk-Factor* was for risk-taking.

When substance use was included as a covariate in the model, the shared genetic loading between *The Addiction-Risk-Factor* and both risk-taking and neuroticism was attenuated down while the association with executive function increased. The reduction in the association with neuroticism is counter to expectations from the stage-based model which posits a more prominent role of negative affect for SUD relative to substance use. We speculate that neuroticism, which represents an amalgam of negative affect traits, may be too broad a construct when considering SUD-specific negative affect; large-scale studies of domains of negative affectivity (e.g., negative urgency) are needed.

### Non-substance Psychopathology and *The Addiction-Risk-Factor*

We found that the 3 non-substance psychopathology clusters, derived from 8 psychiatric disorders^14^, were genetically associated with *The Addiction-Risk-Factor*. The association with early neurodevelopmental disorders, which include ADHD, was the strongest. Cross-loading ADHD on *The Addiction-Risk-Factor* to condition on ADHD attenuated the loading but it remained high. Associations between *The Addiction-Risk-Factor* and the psychopathology clusters were greater than associations with trait representations of behavioral stages of addiction (with the exception of risk-taking). For instance, the genetic association between *The Addiction-Risk-Factor* and the two disorder clusters that included Major Depressive Disorder (i.e., psychotic disorders and early neurodevelopmental disorders) was greater in magnitude than the *Addiction-Risk-Factor*-neuroticism association. Interestingly the compulsive disorder factor did not show strong associations with *The Addiction-Risk-Factor*, suggesting that compulsive disorders and addiction-related compulsive behaviors have distinct etiologies.

Of the 3 behavioral correlates, risk-taking was the most prominent contributor to the association between *The Addiction-Risk-Factor* and all non-substance psychopathology factors. After accounting for substance use, only risk-taking and executive function mediated *The Addiction-Risk-Factor* associations with the psychotic disorder factor. Executive function maintained the only direct association with *The Addiction-Risk-Factor* after accounting for genetics of substance use and genetics of non-substance psychopathology. Thus, we speculate that while risk-taking may characterize the genetic overlap between substance use and other psychopathology, executive function impairment is a risk factor that not only shapes the overlap between addiction and non-substance psychopathology but also explains variance in addiction above and beyond that overlap.

### Limitations

There are several limitations. First, we had to restrict our analyses to individuals of European descent due to the lack of well-powered discovery GWAS informative for other ancestry groups. Second, to maximize sample size of discovery GWASs, our alcohol and tobacco use GWAS incorporated measures of “problematic” use that, while genetically highly correlated with AUD and ND, may include behavioral patterns that are less severe than those represented by use disorder and were not assessed based on clinical presentation. Third, the analyses contain an over-representation of men, in part because the MVP samples contributed most of OUD and half of PAU and the MVP is ∼90% male. Studies with larger numbers of women would allow stratified analyses to explore the differences between sexes observed in epidemiological studies. Fourth, while it is unlikely that individuals completed assessments of risk-taking, neuroticism, and executive function while under the influence of substances, how substance use may have influenced these assessments cannot be determined. Fifth, though significant, mediation pathways were small in effect. Sixth, how these processes effect phenotypic patterns is unknown, however twin studies support a common factor model as well^4^.

## Conclusions

Common genetic liability undergirds distinct SUDs and shares variance with putative behavioral intermediary phenotypes/SUD risk factors and non-substance psychopathology. This addiction genetic factor is more than a linear combination of substance use and psychopathology; it represents a unique addiction dimension that is partially captured by executive functions.

## Data Availability

All data are made publicly available through their respective consortiums.

## Acknowledgments

We gratefully acknowledge our contributing studies and the participants in those studies without whom this effort would not be possible.

The MVP summary statistics were obtained via an approved dbGaP application (<u>phs001672.v4.p1</u>). The authors thank Million Veteran Program (MVP) staff, researchers, and volunteers, who have contributed to MVP, and especially participants who previously served their country in the military and now generously agreed to enroll in the study. (For details, see https://www.research.va.gov/mvp/ and Gaziano, J.M. et al. Million Veteran Program: A mega-biobank to study genetic influences on health and disease. J Clin Epidemiol 70, 214-23 (2016)). This research is based on data from the Million Veteran Program, Office of Research and Development, Veterans Health Administration, and was supported by the Veterans Administration (VA) Cooperative Studies Program (CSP) award #G002.

This study included summary statistics of a genetic study on cannabis use (Pasman et al, 2018 Nature Neuroscience). We would like to acknowledge all participating groups of the International Cannabis Consortium, and in particular the members of the working group including Joelle Pasman, Karin Verweij, Nathan Gillespie, Eske Derks, and Jacqueline Vink. Pasman et al, (2018) included data from the UK Biobank resource under application numbers 9905, 16406 and 25331.

## Supplemental Tables and Figures

### Alternative Model specifications

To establish our factor model, we tried several different approaches. First, we confirmed that the problematic alcohol use (PAU) and problematic tobacco use (PTU) variables did not significantly alter factor structure, as these are more proxies of the underlying construct of interest. Samples that are about an order of magnitude smaller are available for this exploration.

First, we reran the model with alcohol dependence from the Psychiatric Genetics Consortium GWAS of European descent (N= 38,686)^1^ as the indicator for alcohol use disorder, instead of the larger problematic alcohol use (PAU) GWAS (Models M1). As the PGC alcohol dependence GWAS did not include data from the MVP, we also used this as an opportunity to test whether the residual genetic correlation between opioid use disorder (OUD) and alcohol dependence was present in the absence of measurement overlap. The model fit well (*X*^*2*^ = 1.22, p = .54, df = 2, CFI = 1). When we tested the residual correlation, it was large although no longer significant (r=0.71, P=4.46e-01). The residual variance for alcohol dependence was also not significant (r=0.207 p=.51). As the alcohol dependence measure without MVP does not have a significant residual correlation, we posit that the need for the residual correlation when the MVP PAU summary data are used is to improve fit due to the larger sample or due to *measurement* overlap in the MVP.

Second, we tested whether inclusion of cigarettes per day with FTND created a poor proxy for tobacco dependence (Model M2). The model with FTND an indicator fit well, (*X*^*2*^ = 0.027, p = .987, df = 2, CFI = 1) FTND indictor pathway was larger, but the confidence intervals overlapped substantially (*r* = .502, CI = .121).

For our next test of robustness, we conducted two follow-up analysis to examine whether parsing *Addiction-rf* into a two-factor structure improved fit (Models M3). The highest pairwise associations between SUD indicators were OUD with PAU. PTU with CUD also showed a high pairwise associations. Both factors were allowed to correlate. This model did not fit the data well (model fit: *X*^*2*^ = 34.039, p = 1.94e-07, df = 3, CFI = .924, AIC = 47.039). We also fit a model where PAU and PTU loaded on one factor (“licit drugs”) while CUD and OUD loaded on another correlated factor (“illicit drugs”). This model did not fit the data well either (model fit: *X*^*2*^ = 293924.2, p = <.001, df = 5, CFI = -721.5134, AIC = 293934.2).

Next, there are two alternative models we tested to see if there was a more appropriate analysis to account for substance use (Models M4). To be clear, substance use is a covariate, but its covariate effect may be better modeled via alternative models. First, we generated a substance use factor using ever smoking regularly, marijuana use, and drinks per week as indicators. We then regressed the use disorder indicators on this factor. This model fit the data poorly. (*X*^*2*^*=*394.4224, df=1, p=5.739041e-77 AIC=426.4224 CFI=0.8833732) and significantly worse than our original model (*X*^*2*^ *diff=*394.007, df=9, p=2e-64).

Next, rather than have the substance use variables endogenous to the indicators of addiction factor, we tested whether an alternative model where the indicators predicted the addiction factor directly fit the data better via a nested comparison. A model with the substance use variables predicting addition-*rf* fit the data poorly (*X*^*2*^=324.481, df=10, P=1.025754e-63, AIC=360.481, CFI=0.9040932).

### Q-Trait Analysis

#### Background. Background

Q-Trait analysis^1^ was developed to test whether a latent genetic factor estimated using gSEM mediates the genetic association between an external trait (i.e., one not loaded onto the latent genetic factor) and factor indicators (those loading on the latent genetic factor). We conducted Q-trait analysis to test whether The *Addiction-Risk-Factor* accounts for genetic associations between these traits and each individual SUD indicator loading onto it (i.e., OUD, PAU, PTU, CUD). Q-Trait analysis evaluates this by comparing one model in which only the latent genetic factor is associated with the external trait to another model in which each indicator (i.e., the trait loading onto the genetic factor) is associated with the external trait. A *χ*2 difference between these models is calculated and if there is a significant difference, it suggests that the genetic association between the external trait and the factor indicators are not accounted for by the latent genetic factor. However, this analysis is conducted with all factor indicators in a single model independent of the latent factor, and does not model scenarios in which the latent factor may differentially mediate (e.g., fully, partially, or not at all) individual factor indicators. Because of this limitation, in cases where we found no evidence that the latent genetic factor completely mediated the genetic association between the external trait and factor indicators (i.e., when there was a significant *χ*2 difference), we conducted a series of post-hoc models regressing the latent genetic factor and each individual genetic indicator on the external traits in an iterative processes (For example, a model in which the latent genetic *Addiction-Risk-Factor* and problematic tobacco use were regressed on executive function).

#### Results

Q-Trait analysis revealed that the latent genetic *Addiction-Risk-Factor* mediated associations between risk-taking and individual SUD indicators, as indicated by a lack of significance in model fit (*X*^*2*^*diff(3) =* 6.478, P=0.091). Although initial Q-trait modeling provided no evidence that the latent genetic Addiction-Risk-Factor accounted for associations between individual SUD indicators with executive function (*X*^*2*^*diff(3) =* 25.7, P = 1.00e-5) and neuroticism (*X*^*2*^*diff(3) =* 625.7, P=1.00e-5), post-hoc analyses revealed that the lack of evidence for mediation is attributable to differential mediation across indicators. Executive function is correlated with *The Addiction-Risk-Factor*, but shows stronger associations with problematic alcohol use, cannabis use disorder, and opioid use disorder than problematic tobacco use (see **Supplemental Table 1**). Because of this we tested an additional model wherein the latent genetic *Addiction-Risk-Factor* and problematic tobacco use were regressed on executive function simultaneously, which revealed no significant difference from the model in which all individual SUD indicators were included (*X*^*2*^*diff(2) =*4.635, P = .0985), suggesting that the common latent *Addiction-Risk-Factor* mediates associations between executive function and PAU, OUD, and CUD, but not PTU. Further, additional regression path greatly bolstered upward the association *between The Addiction-Risk-Factor* and executive function (r=-.31, p = 1.00e-17). For neuroticism, the addition of specific paths with PAU and CUD in addition to *The Addiction-Risk-Factor* revealed evidence that the latent genetic factor mediates associations with OUD and PTU (*X*^*2*^*diff(1) =* 2.798, P=0.0944). However, this did not change the association with the latent factor (r=.20, p=7.08e-3); it is simply that neuroticism has associations with PAU and CUD that are independent of *The Addiction-Risk-Factor*.

#### Interpretation for downstream analysis

Downstream analysis did not retain these specific associations. For executive functioning, we still found significant associations between cEF and addiction risk, and mediating pathways despite the association being attenuated downward by our chosen model. Neuroticism did not maintain mediating pathways, and so improving the model would not have led to additional discovery power for the neuroticism phenotype.

**Table 1.**
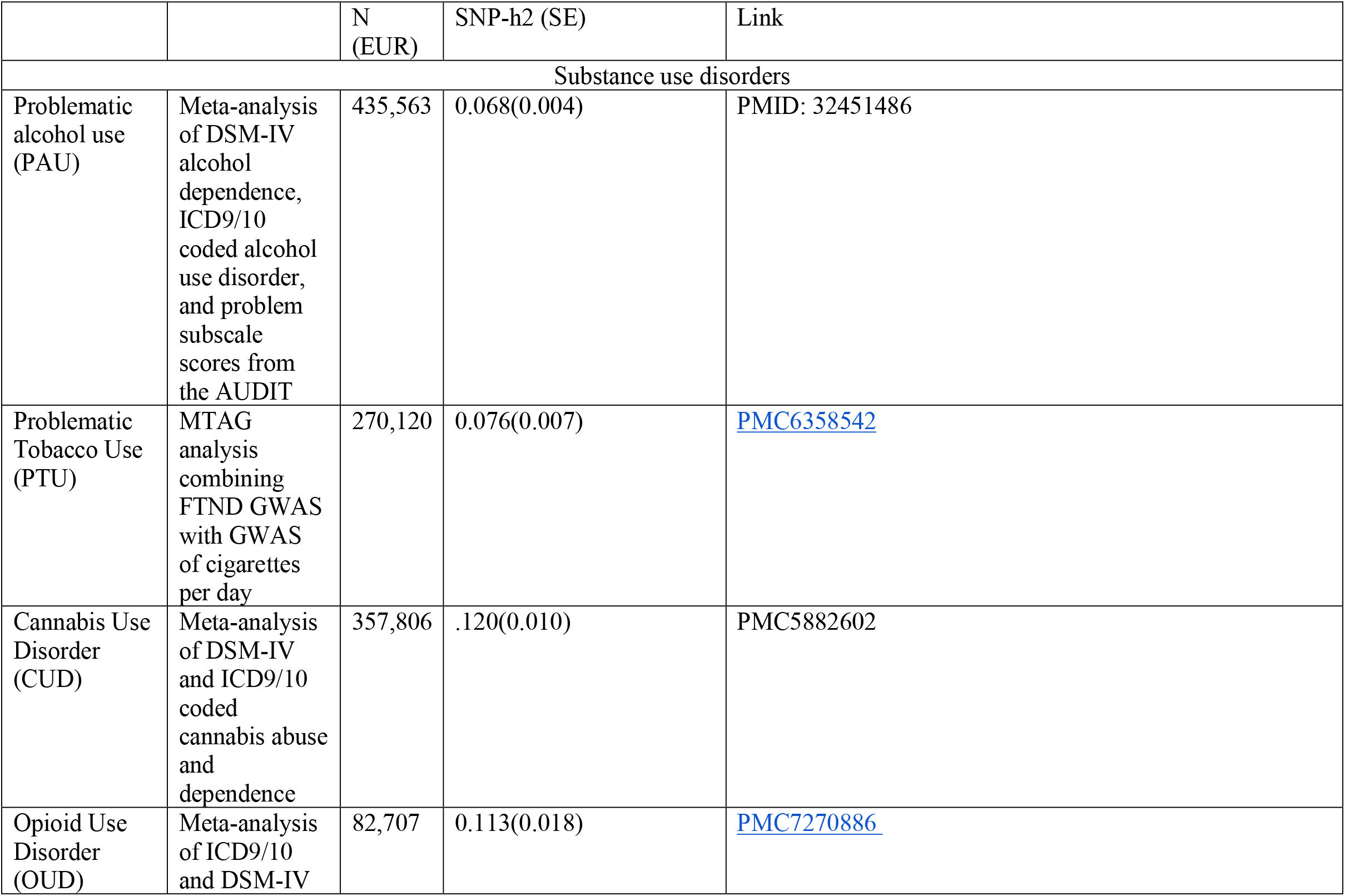
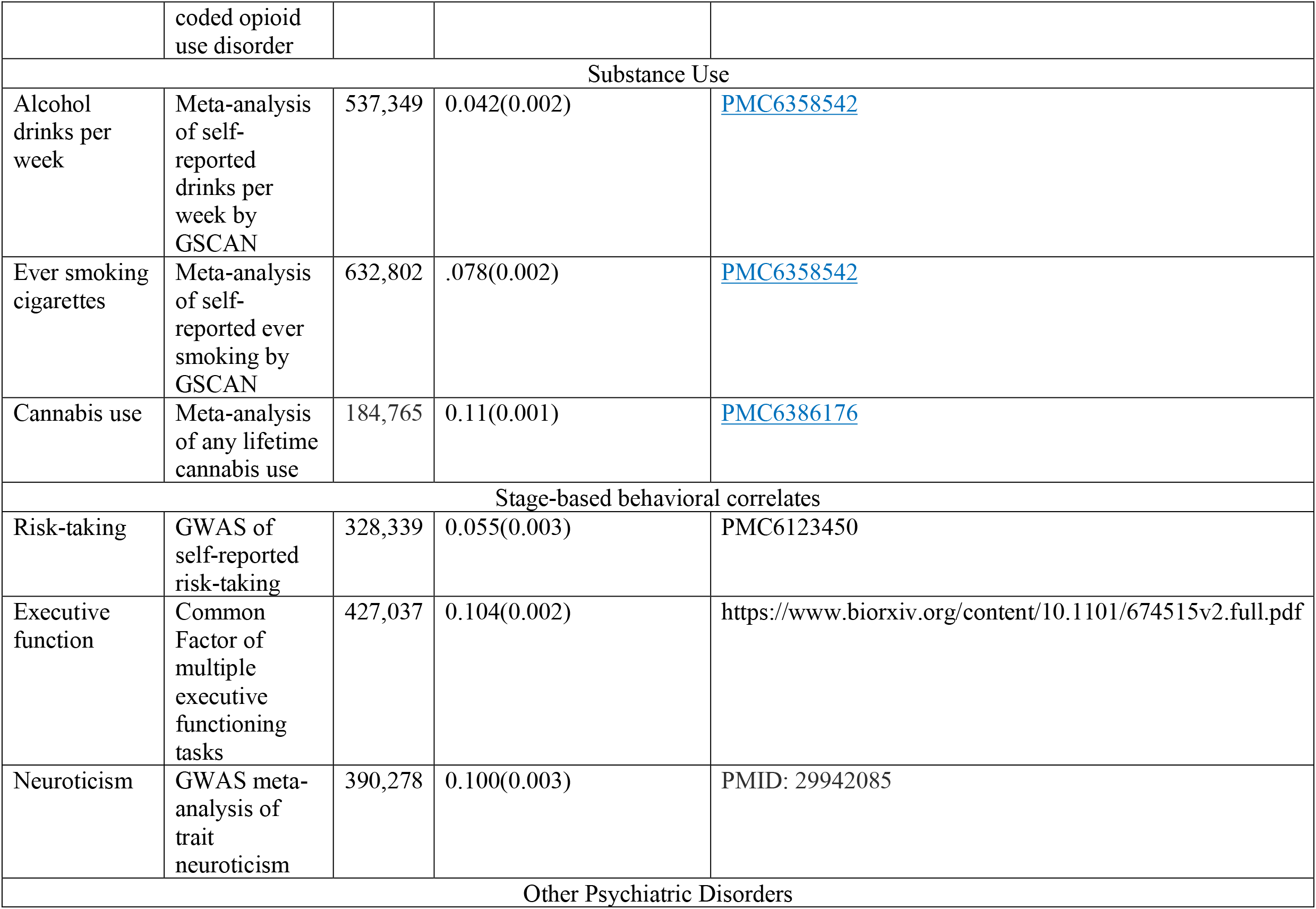
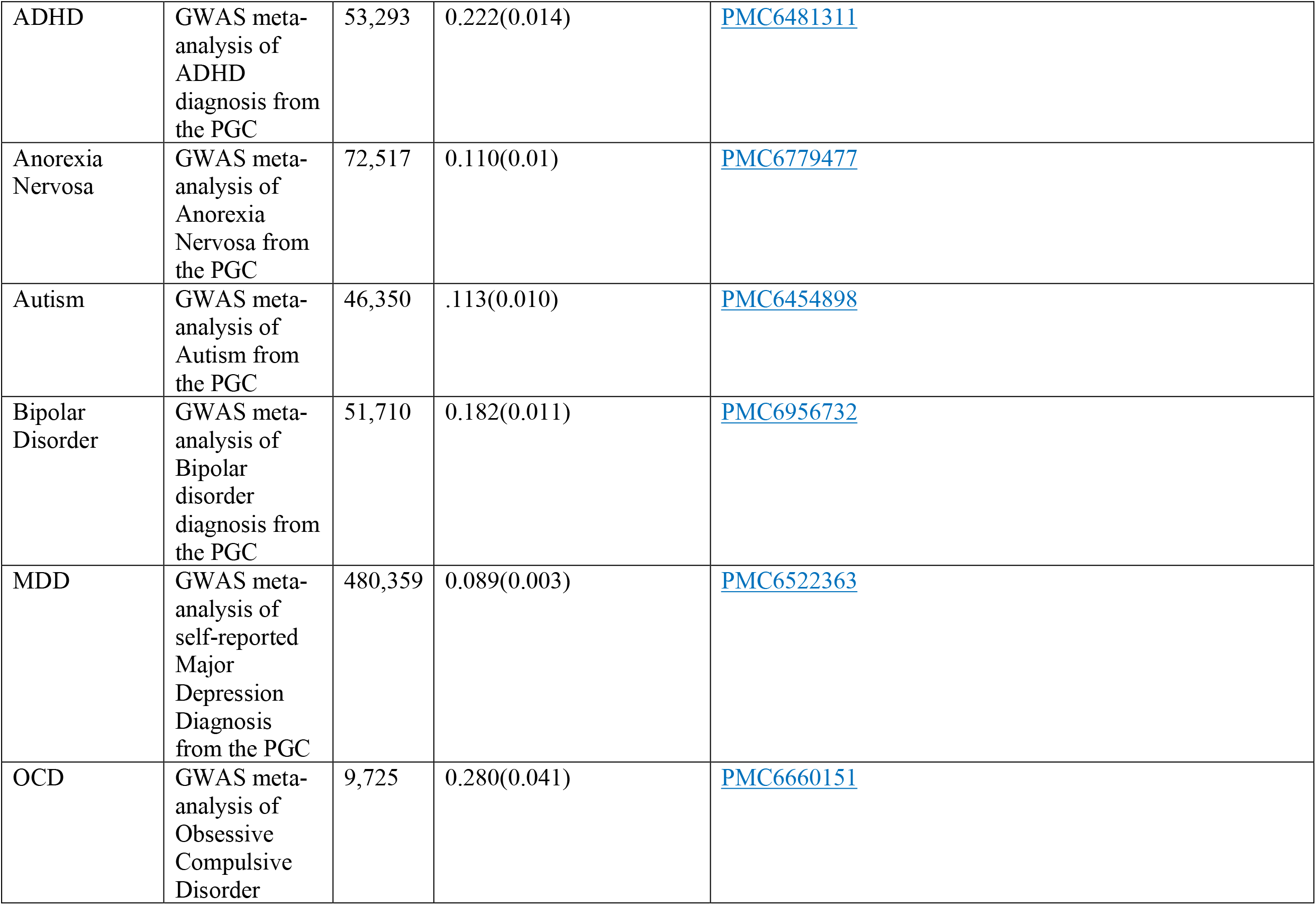
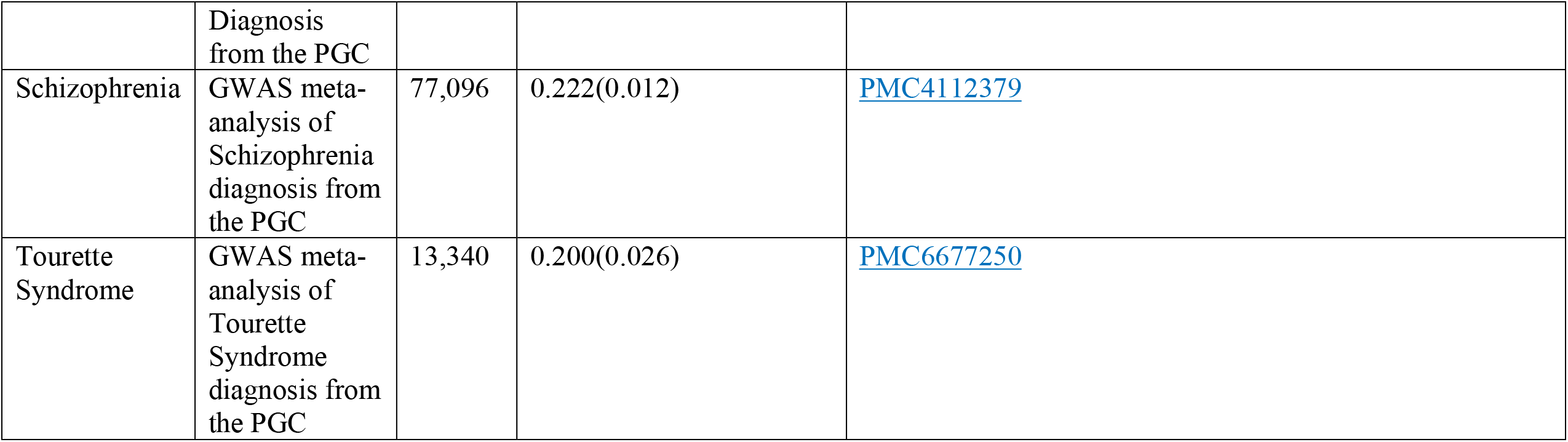
GWAS summary statistics used in this study.

**Supplementary Table 2.**
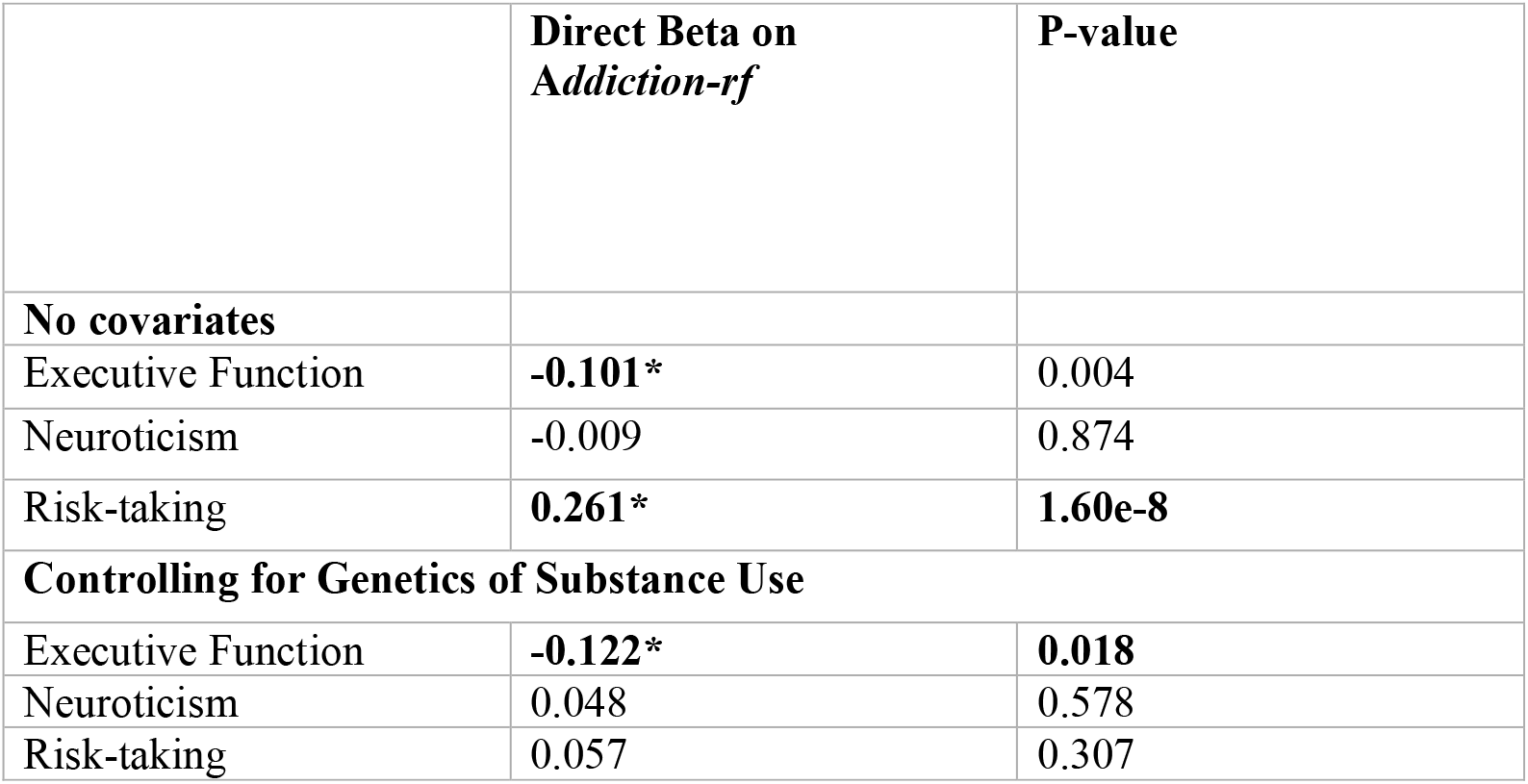
Direct Associations between behavioral correlates and the Addiction Risk Factor – *Addiction-rf* - beyond non-substance psychopathology. **Behavioral Liabilities Mediate the Association Between -Factor and Psychiatric Factors** Indirect associations from a mediation model (see Figure 4) where stage-based constructs link non-substance psychopathology (three factors from Lee et al.,) and *The Addiction-Risk Factor* (*Addiction-rf)*. Indirect association is represented by the proportion accounted for by the indirect association (%) and the significance of the indirect association. F1 = compulsive disorders, F2 = Psychotic disorders, F3 = Neurodevelopmental disorders.

**Supplementary Figure 1.**
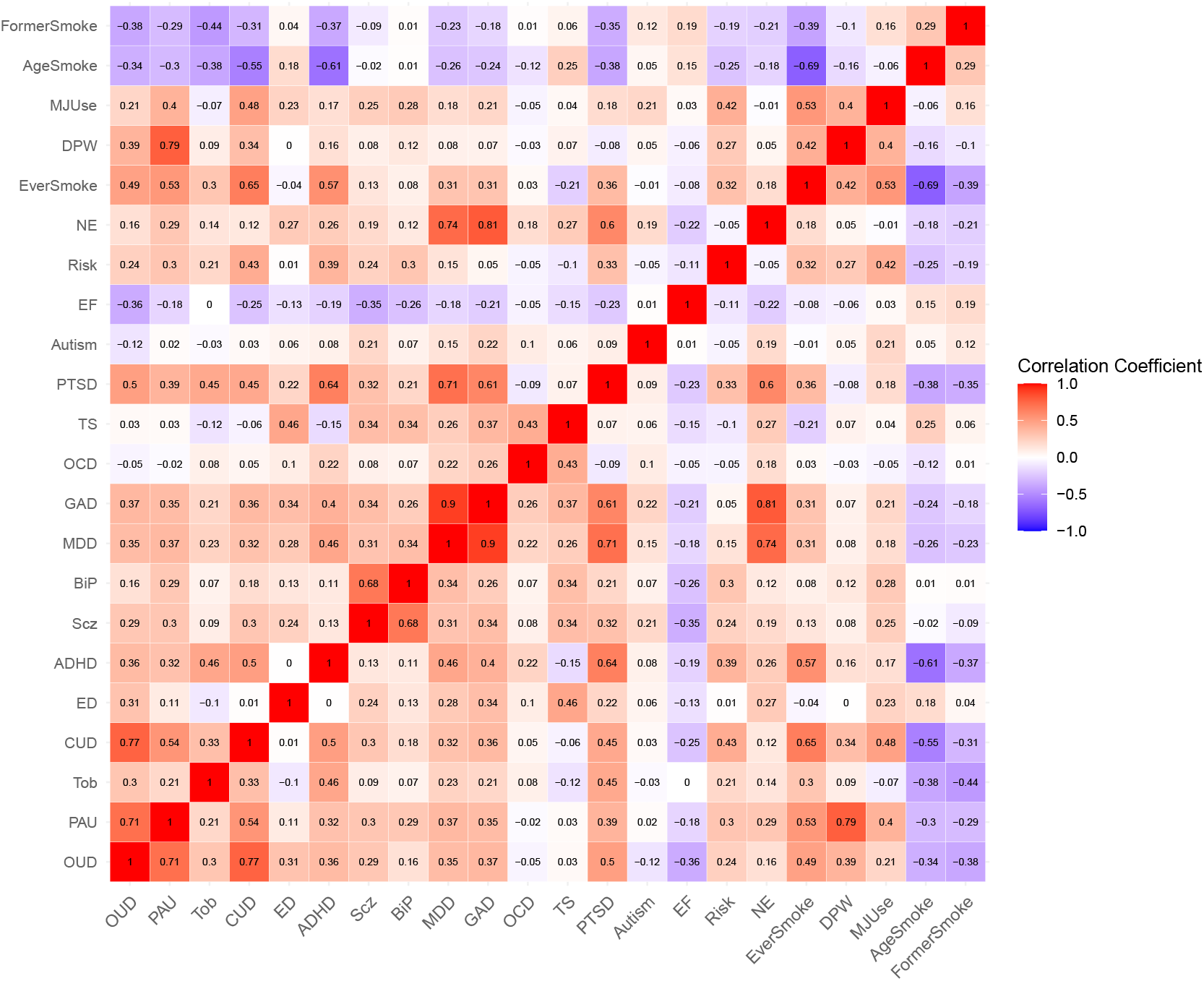
Genetic correlations (SNP-rG) were estimated between all variables. The upper diagonal represents the bivariate associations from LD Score regression. PAU = Problematic Alcohol Use, OUD = Opioid Use Disorder CUD = Cannabis Use Disorder. PTU = Problematic Tobacco Use. FormerSmoke = smoking cessation, AgeSmoke=Age first smoking regularly, MJUse = Marijuana Use, DPW = Drinks Per Week, EverSmoke = Ever Smoking regularly, NE = neuroticism, Risk = Risk-taking, EF = Executive Function, PTSD = Post-Traumatic Stress Disorder, TS = Tourette’s Syndrome, OCD = Obsessive Compulsive Disorder, GAD = General Anxiety Disorder, MDD = Major Depressive Disorder, BiP = Bipolar disorder, Scz = Schizophrenia, ADHD =

**Supplementary Figure 2.**
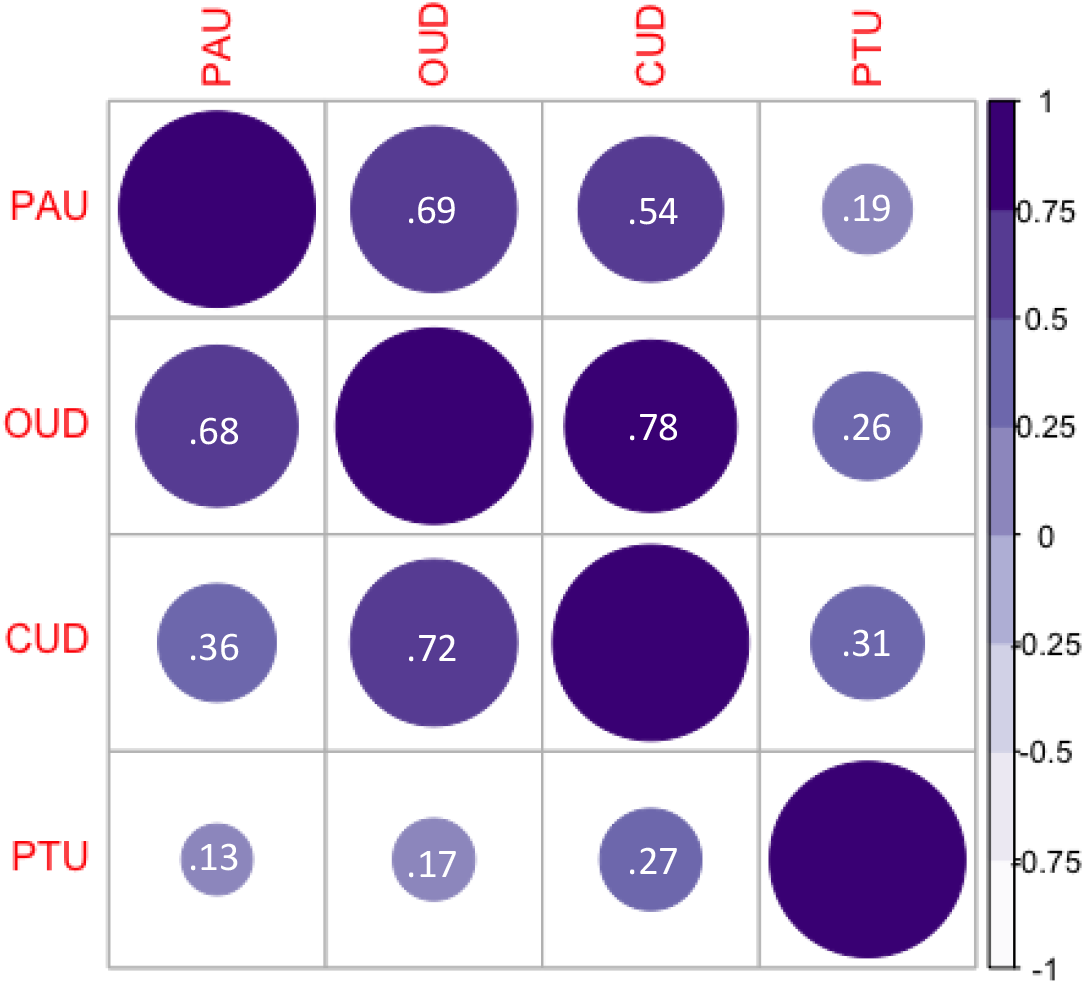
Genetic Correlation Matrix Between Substance Use Disorders. Genetic correlations (SNP-rG) were estimated between all substance use disorder categories. The upper diagonal represents the bivariate associations from LD Score regression. The lower diagonal are correlations estimated in Genomic Structural Equation Modeling when controlling for common substance use (drinks per week, ever smoking marijuana or tobacco). All genetic correlations were positive (shown in purple). PAU = Problematic Alcohol Use, OUD = Opioid Use Disorder CUD = Cannabis Use Disorder. PTU = Problematic Tobacco Use. All rGs were significant (all p> .01).

**Supplementary Figure 3.**
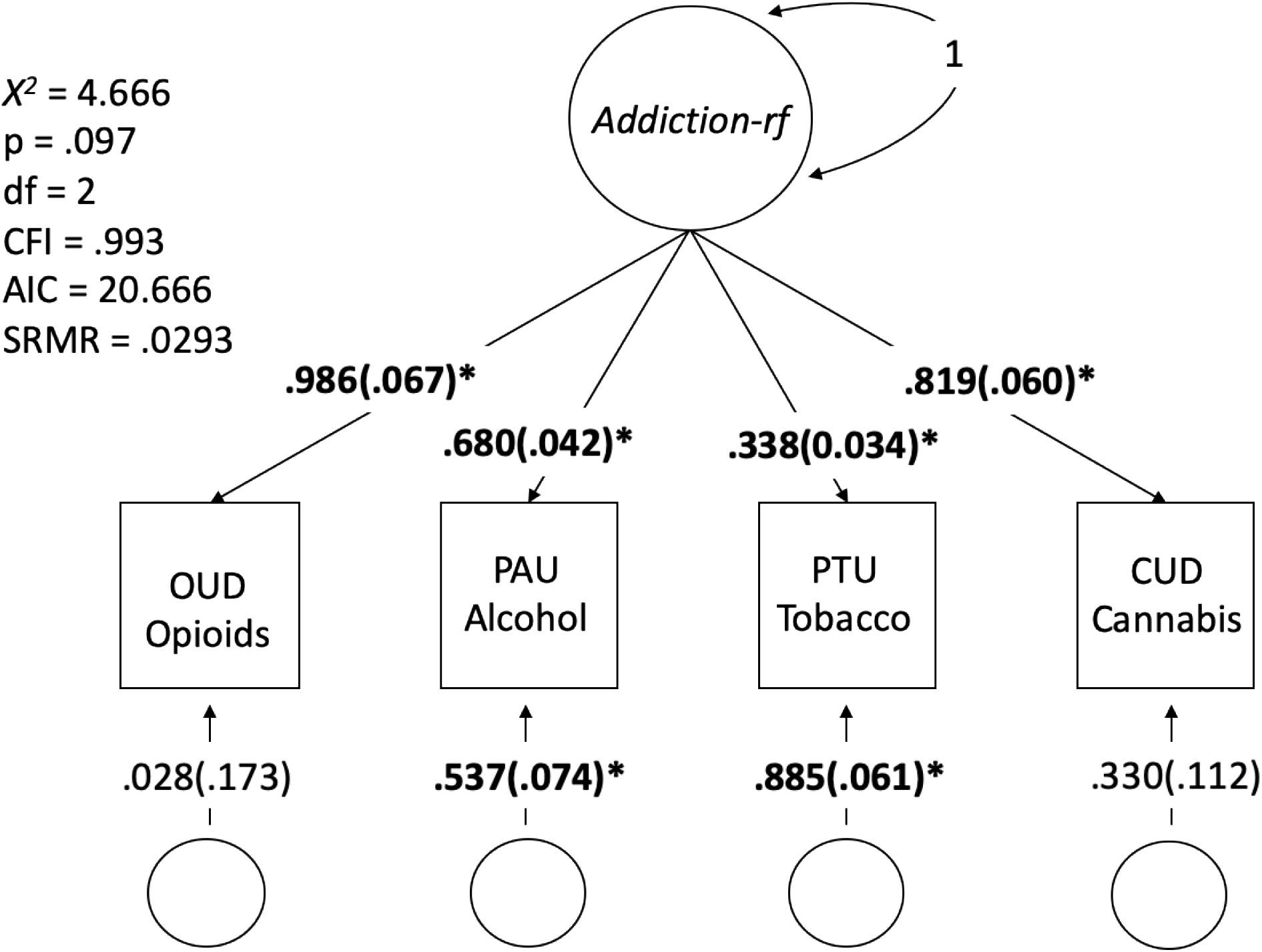
Alternative specification of one factor model, without residual correlation between problematic alcohol use and opioid use disorder.

**Supplemental Figure 4.**
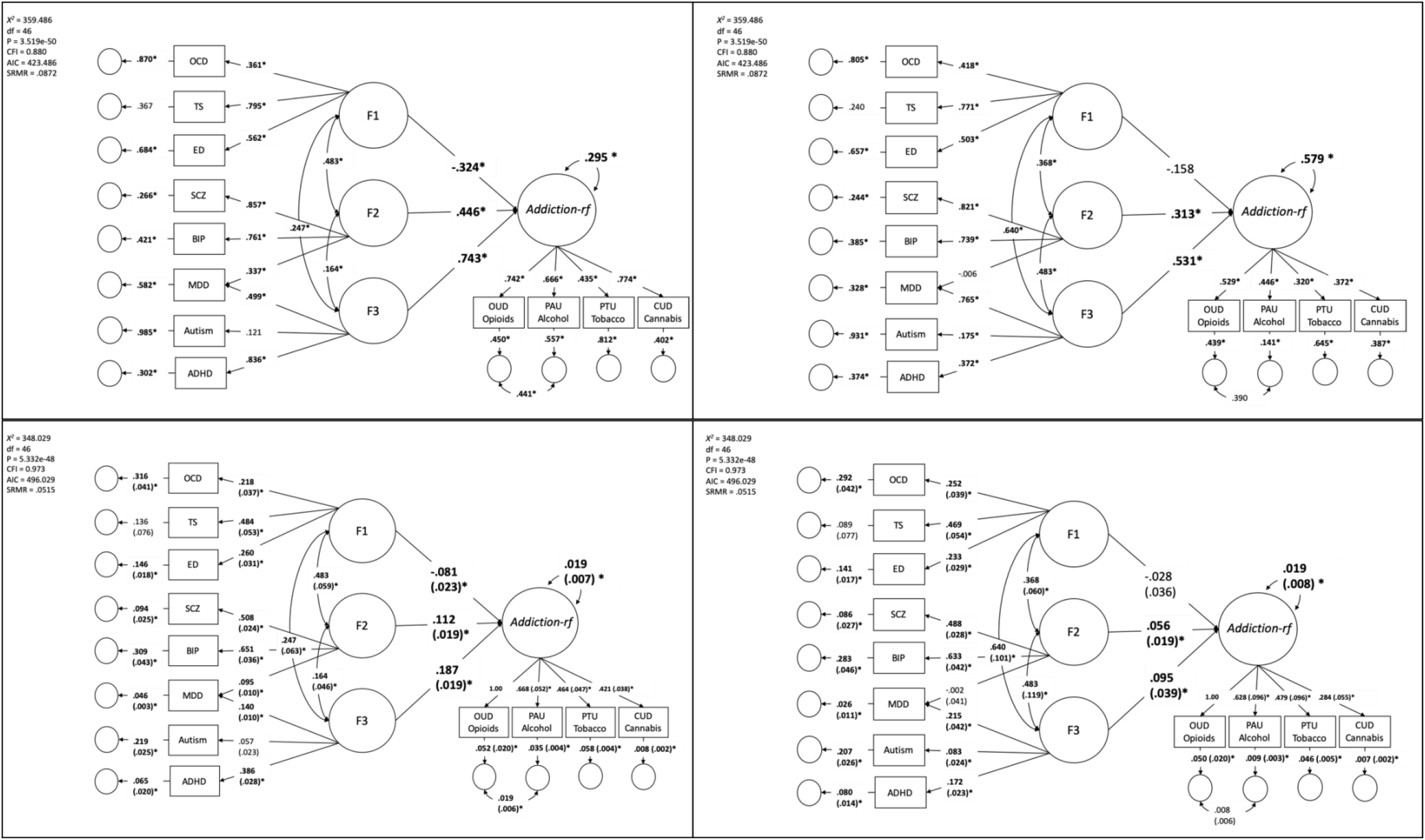
Full model specification for *Addiction-rf* association with non-substance psychopathology. The top two panels represent the standardized estimates, the bottom two are unstandardized with standard errors. The left panels include the substance use variables as covariates. **Bold*= p-value < .05**.

**Supplementary Figure 5.**
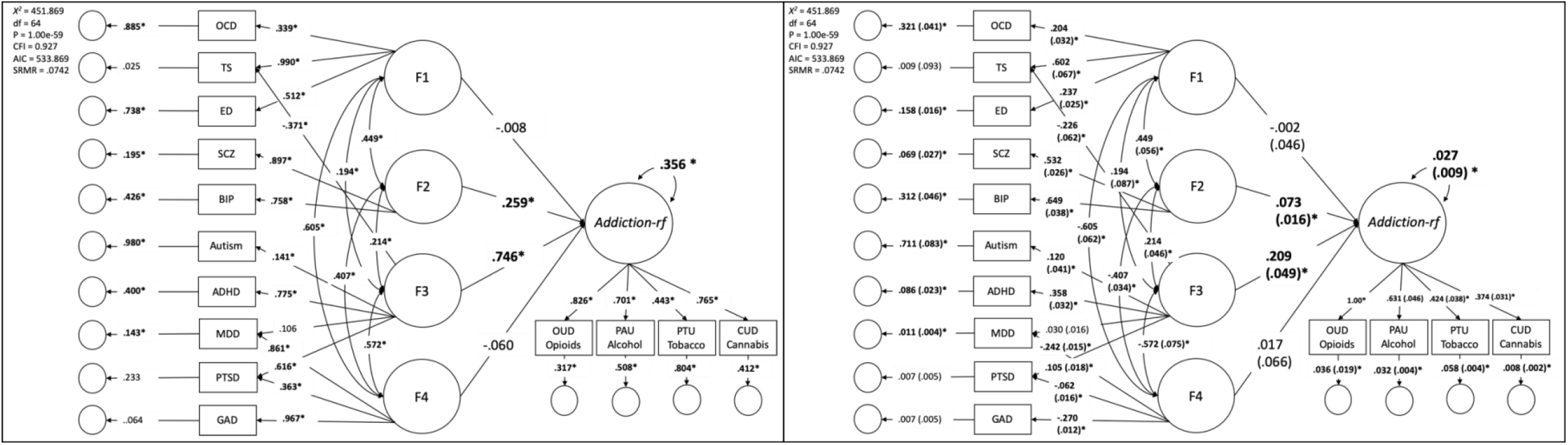
Alternative model of psychopathology, taken from Grotzinger et al.^2^ the left panel represents the model with standardized estimates. The right panel represents the unstandardized estimates with standard errors. **Bold*= p-value < .05**.

**Supplemental Figure 6.**
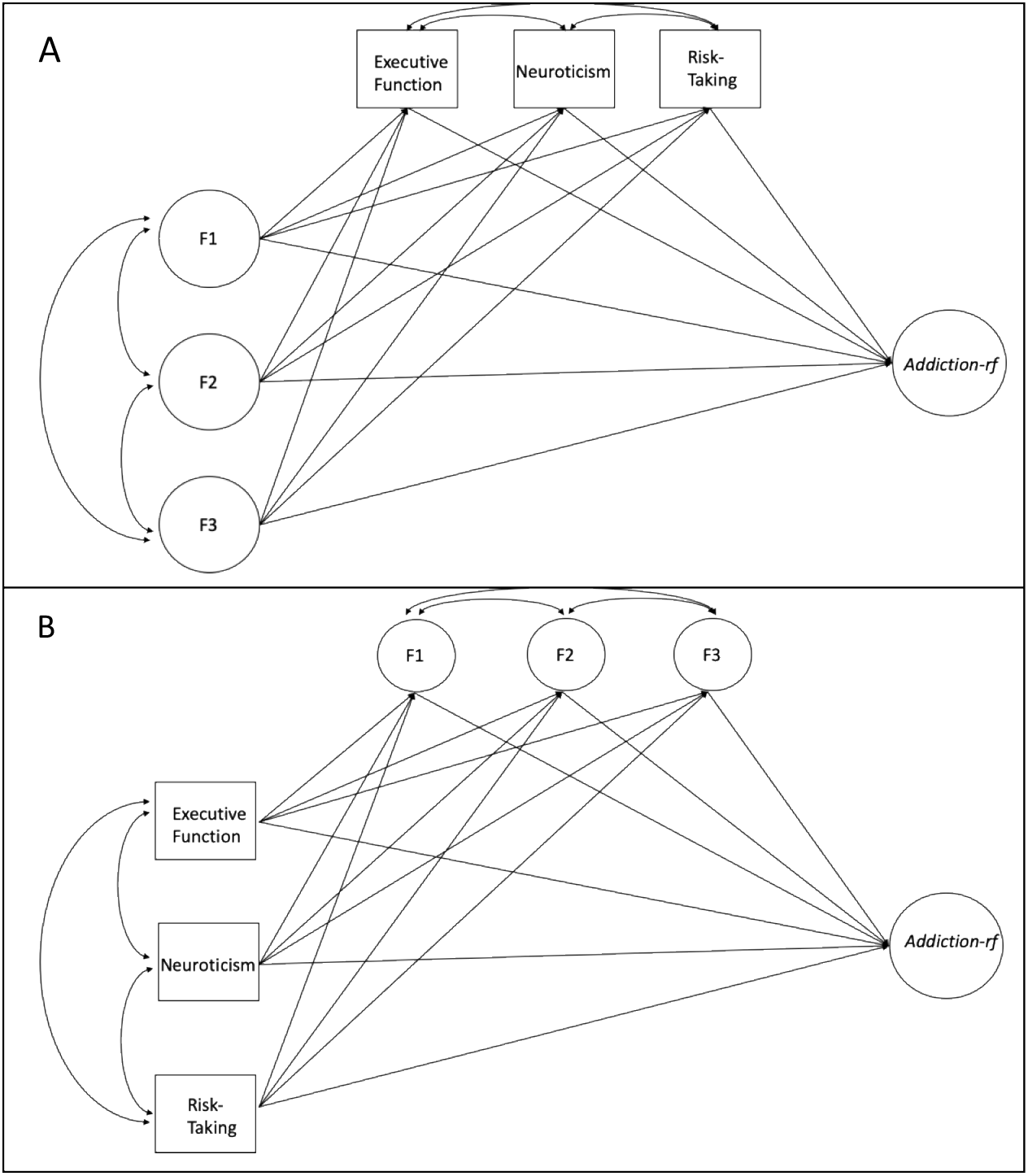
Model Specification for Mediation Models Linking non-substance psychopathology and *Addiction-rf*. In panel A, we tested whether neurobiological stage-based constructs mediated the association between the *Addiction-rf*-Factor and non-substance psychopathology. In mediation model two, we tested whether neurobiological stage-based constructs had remaining (direct) associations with *Addiction-rf* beyond non-substance psychopathology variables.

